# Revisiting the concept of “phenotype” in pediatric hypertrophic cardiomyopathy using myocardial stiffness and strain variations assessed by ultrafast ultrasound imaging

**DOI:** 10.1101/2025.11.04.25339540

**Authors:** Aimen Malik, Maelys Venet, Seema Mital, Clement Papadacci, Mathieu Pernot, Mark Friedberg, Luc Mertens, Jerome Baranger, Olivier Villemain

## Abstract

**Background:** Pediatric hypertrophic cardiomyopathy (HCM) is associated with significant morbidity and mortality. While identified as a genetic disease mainly involving sarcomeric genes, the association between genotypical variation and phenotypic expression is not fully established. Developments in Ultrafast ultrasound imaging allows quantifying myocardial stiffness using shear waves elastography (SWE). When combined with strain measurements, myocardial work can be computed, offering new insights into phenotype myocardial properties.

**Methods:** An age-matched population of 20 Healthy volunteers (HVs, mean age=11.1 ± 4.5years), 20 HCM (genotype- and phenotype-positive, mean age=11.6 ± 5.3years) and 20 Genotype (genotype-positive, phenotype-negative, mean age=11.1 ± 4.8years) were included in the study. Each participant underwent conventional echocardiography and a full cardiac-cycle exploration of the basal anteroseptal segment consisting of: (1) myocardial stiffness by SWE, (2) segmental strain and thickness, which are used to compute one-beat work, the stress-strain loop area, contributive and dissipative work.

**Results:** Mean diastolic myocardial stiffness (DMS) and peak myocardial strain (PMS) distinguished the HCM group (DMS=23.7 ± 8.7kPa; PMS= −6.64 ± 5.9%) from HV group (DMS=7.2 ± 0.7kPa, p<0.01; PMS= −19.9 ±4.1%, p<0.01). No significant differences were observed in DMS and PMS between HVs and Genotype groups. One-beat work and stress-strain loop areas showed significant differences among all 3 groups (p<0.01) and could distinguish the Genotype group (one-beat work= 318.2 ± 100.2µJ/mm; stress-strain loop area=33.2 ± 10.6kPa.%) from HVs (one-beat work= 582.4 ± 137µJ/mm; stress-strain loop area= 66.8 ± 19.8 kPa.%), and the HCM group (one-beat work= 38.2 ± 107.1µJ/mm, stress-strain loop area=5.8 ± 6.5kPa.%), p<0.01.

**Conclusion:** Combining Ultrafast ultrasound with speckle-tracking echocardiography, we demonstrate that one-beat work and stress-strain relationship, obtained by combining myocardial stiffness, strain, and thickness have the potential to distinguish genotype-positive, phenotype-negative patients from healthy controls. Clinical outcome studies are needed to determine the prognostic value of these parameters in phenotype-positive HCM patients.

## INTRODUCTION

Hypertrophic cardiomyopathy (HCM) is the most common inherited cardiac disease and is the leading cause of sudden cardiac death in children and young adults[1]. HCM is currently diagnosed by the presence of left ventricular hypertrophy (LVH) unexplained by another cardiac, systemic or metabolic disease. While sarcomere gene mutations are identified as the most prevalent cause of HCM, the phenotypic expression is extremely heterogeneous ranging from patients with early disease progression resulting in severe hypertrophy at high-risk for developing end-stage heart failure or sudden cardiac death [1], [2], [3], [4]. However, the link between genotype and cardiac phenotype in HCM patients remains unclear and current risk prediction models have got significant limitations [5], [6], [7]. As this is a primary myocardial disease, imaging techniques that could help to better characterize myocardial properties in this high-risk population could help to better define the cardiac phenotype and could potentially help to identify high-risk patients.[8].

It has been shown that development of systolic dysfunction is a strong predictor of major adverse cardiac effects in HCM patients, but the disease process can also affect diastolic function with diastolic abnormalities been described even before LVH develops [1] [9]. Moreover, in patients with significant LVH, development of diastolic dysfunction negatively impacts outcomes. Unfortunately, identification of diastolic dysfunction based on conventional echocardiographic parameters can be challenging in children with cardiomyopathy [10].

The introduction of ultrafast ultrasound and the application of myocardial shear wave elastography (SWE), allows to estimate changes in myocardial stiffness (MS) throughout the cardiac cycle non-invasively [11]. We previously demonstrated increased diastolic myocardial stiffness in adult HCM patients compared to healthy controls [12], [13], [14], [15]. In this study we want to further characterize pediatric HCM phenotype based on myocardial stiffness analysis and, by combining stiffness assessment with strain and thickness assessment throughout the cardiac cycle, it becomes possible to calculate myocardial work and the stress-strain relationship throughout the cardiac cycle. We aim to study this in pediatric HCM patients with sarcomeric mutations presenting with different phenotypes, including those with no hypertrophy (genotype positive, phenotype negative). We hypothesized that detailed myocardial phenotyping by these methods may detect differences in myocardial properties early in the disease process that may be difficult to detect using conventional echocardiographic parameters.

## METHODS

### Population and design

We prospectively enrolled three age-matched groups of participants (0-18 years of age) at The Hospital for Sick Children, Toronto, Ontario, Canada: (1) HCM patients (genotype-positive, phenotype-positive), (2) Genotype patients (genotype-positive, phenotype-negative), and (3) healthy volunteers (HVs). The inclusion criteria for the HCM group were the presence of LV hypertrophy (LVH) defined as a Z-score of interventricular septum (IVS) ≥ +2.5 in the absence of family history, or ≥ +2 in case of family history, with morphological or genetic evidence for sarcomeric HCM and exclusion of other pediatric HCM causes including secondary hypertrophy from hypertension (HTN), metabolic disorders, or syndromes such as Noonan and LEOPARD, were excluded to ensure a sarcomeric etiology. [1]. The inclusion criteria for the Genotype group was based on the identification of a known genetic mutation that causes sarcomeric HCM according to the 2024 AHA/ACC/AMSSM/HRS/PACES/SCMR Guideline [1]. Inclusion criteria for the HV group were good general health status, absence of current or previous history of congenital or acquired heart disease, and normal echocardiogram. All subjects gave informed written consent, and the study was approved by The Hospital for Sick Children Research Ethics Board (REB # 1000074747).

### Conventional echocardiography

#### Equipment

A full echocardiographic study was performed using GE Vivid-E95 Ultrasound system (GE Healthcare, USA) equipped with a 6S-D or M5Sc-D phased-array probe.

#### Thickness measurement

The thickness of the basal antero-septal segment was measured on the parasternal short axis M-Mode acquisition, throughout the cardiac cycle. During the off-line analysis, the septal thickness values were reported at forty different timepoints of the ECG cycle using a semi-automatized graphic user interface, custom-created in MATLAB software (California, USA), and according to the same ECG-based timescale than the other parameters. The one-beat thickness vs. time curve was thus drawn for each patient and used as is for the one-beat work calculation step.

#### Segmental strain measurement

The myocardial strain of the basal antero-septal segment was obtained on the 3-chamber STE acquisition, throughout the cardiac cycle using GE Ecopac, V202. The offline postprocessing was performed to extract the strain values at forty different timepoints of the ECG cycle using a semi-automatized graphic user interface, custom-created in MATLAB software (California, USA) and according to the same ECG-based timescale than the other parameters. The one-beat strain vs. time curve was thus drawn for each patient and used as is for the one-beat work calculation step (see Figure 1).

**Figure 1.**
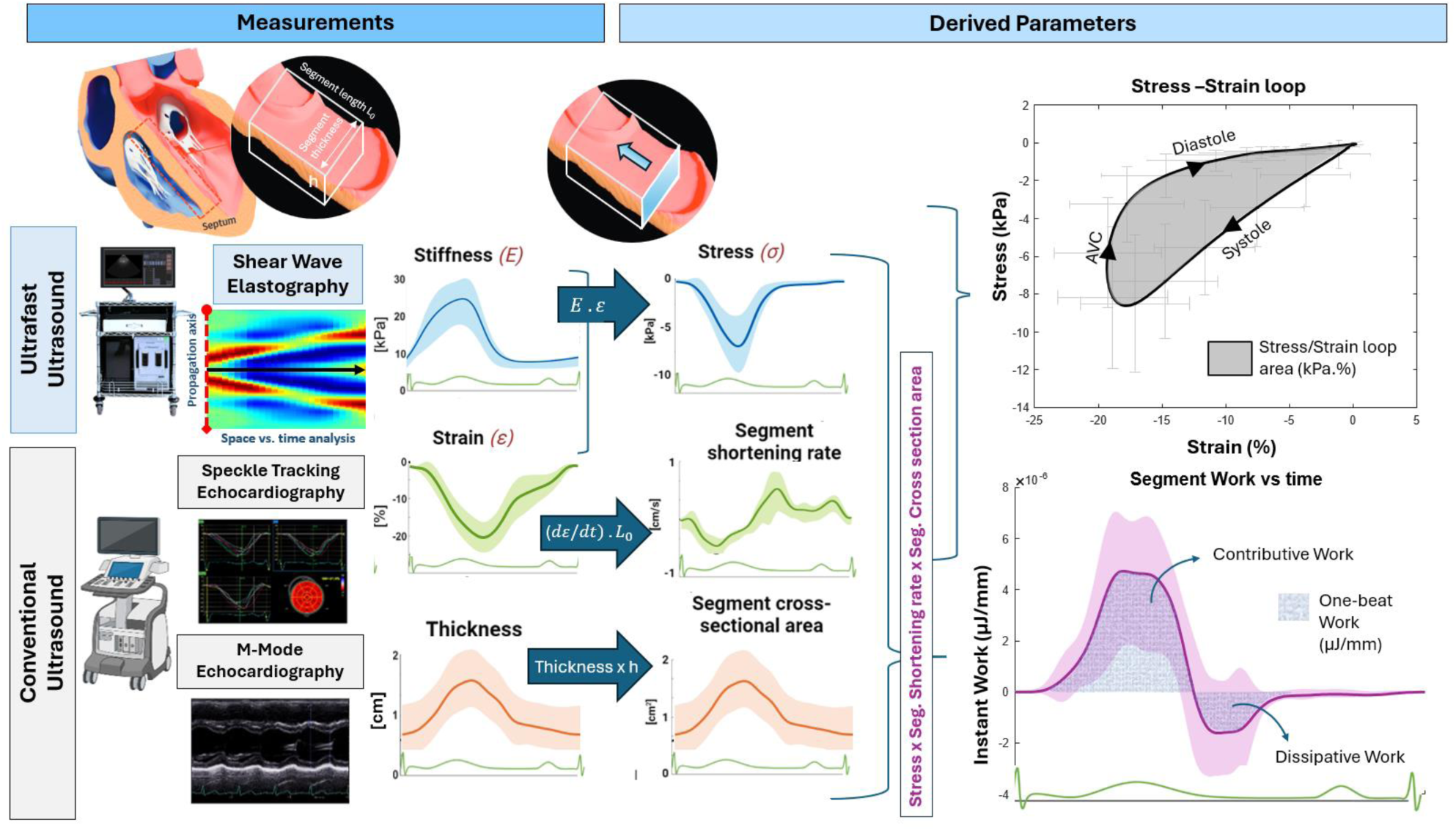
Computing One-beat work and Stress-Strain loops from stiffness, strain and thickness measurements. **Legend**. This figure illustrates the process of computing one-beat work and stress-strain loops using ultrafast and conventional ultrasound techniques. Measurements of stiffness, strain, and thickness are derived from shear wave elastography, speckle tracking echocardiography, and M-mode echocardiography. These parameters are used to calculate myocardial stress, segment shortening, and cross-sectional area, which contribute to the stress-strain loop and segment work analysis, providing insights into cardiac mechanics and efficiency.

#### Clinical HCM genotype information

For the purpose of analyzing genotype-phenotype relationships, the specific gene mutations were classified into two categories: thick myofilament protein mutations (including MYH7 and MYBPC3) and thin myofilament protein mutations (including TNNT2, TNNC1, TNNI3, TTN, and TPM1) in the Genotype group [22]. This classification allows for comparison of potential differences in myocardial mechanics between these mutation types [16].

### Myocardial stiffness assessment by shear wave cardiac elastography

The SWE methods used in this study has previously been described [11], [12], [13], [14], [15], [17]. Ultrafast acquisitions for SWE were performed using the Verasonics Vantage systems (Vantage 256, Verasonics Inc., Kirkland, Washington) with a phased array ultrasound probe (GE 6S-D or M5Sc-D). Briefly, acoustic radiation force induced by a focused ultrasound beam was used to generate shear waves in the myocardial tissue and its propagation velocity was calculated. Shear wave velocities (SWV) were measured in the anteroseptal basal segment of the heart in two orthogonal parasternal views (short and long axis). Post-processing of the SW Imaging data was performed in MATLAB (R2019a, The MathWorks Inc., Natick, MA, USA) and a mean SWV with standard deviation (SD) was calculated for every time point in the cardiac cycle.

Acquisitions throughout the cardiac cycle were obtained using 2 sets of 10 pushes triggered by an electrocardiogram (ECG) with a 100ms incremental delay in parasternal short and long axis views (total of 20 acquisitions in short axis and 20 acquisitions in long axis per participant). The collected acquisitions were then post-processed to visualize the shear waves and compute their velocities [12], [13], [14], [15]. The entire cardiac cycle was divided into 20 time points using ECG wave as a reference with 8 time points in systole and 12 time points in diastole. The ECG trace of every patient was divided into 20 time points using a semi-automatized software to standardize and correct variations related to heart rate and RR interval. Measurements during the isovolumetric contraction (IVC) and isovolumetric relaxation (IVR) periods were obtained at the time of mitral valve closure (MVC) and aortic valve closure (AVC) based on ultrafast B-mode cineloops. Acoustic output strictly complied with the FDA Track 3 recommendations (MI<1.9, TI<3, ISPTA<720mW/cm2 and ISPPA<190W/cm2)[18].

According to the Young modulus, we assessed the myocardial stiffness (E) using the equation:

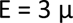

with µ equivalent to shear modulus (µ = ρ. SWV^2^ where ρ is the volumetric mass of the tissue, here equal to 1000kg/m^3^) and assuming that myocardium Poisson’s ratio approaches the value ν = 0.5.

### Myocardial stress and work assessment (Figure 1, Table 1)

Myocardial stress was computed as the product of stiffness and strain over time. Myocardial strain rate was computed by taking the derivative of myocardial stain with respect to time. We define one-beat segmental myocardial work (One-beat work, µJ/mm), as the cumulative work performed by the basal antero-septal segment throughout a cardiac cycle. Essentially, one-beat work quantifies the energy expended by this segment during its contribution to the heart’s contraction. To derive this measure, we assess the energy consumption of the segment at each moment of the cardiac cycle, known as the instantaneous segmental work (µJ/mm). At any given time, one-beat work is defined as the product of the segmental stress, the cross-sectional area upon which this stress acts, and the change in segment length in response to this stress. These parameters can be obtained using ultrasound techniques: shear-wave elastography, M-Mode imaging, and strain imaging, respectively. Subsequently, the one-beat work is determined by integrating the instantaneous segmental work over the entire cardiac cycle.

**Table 1.**
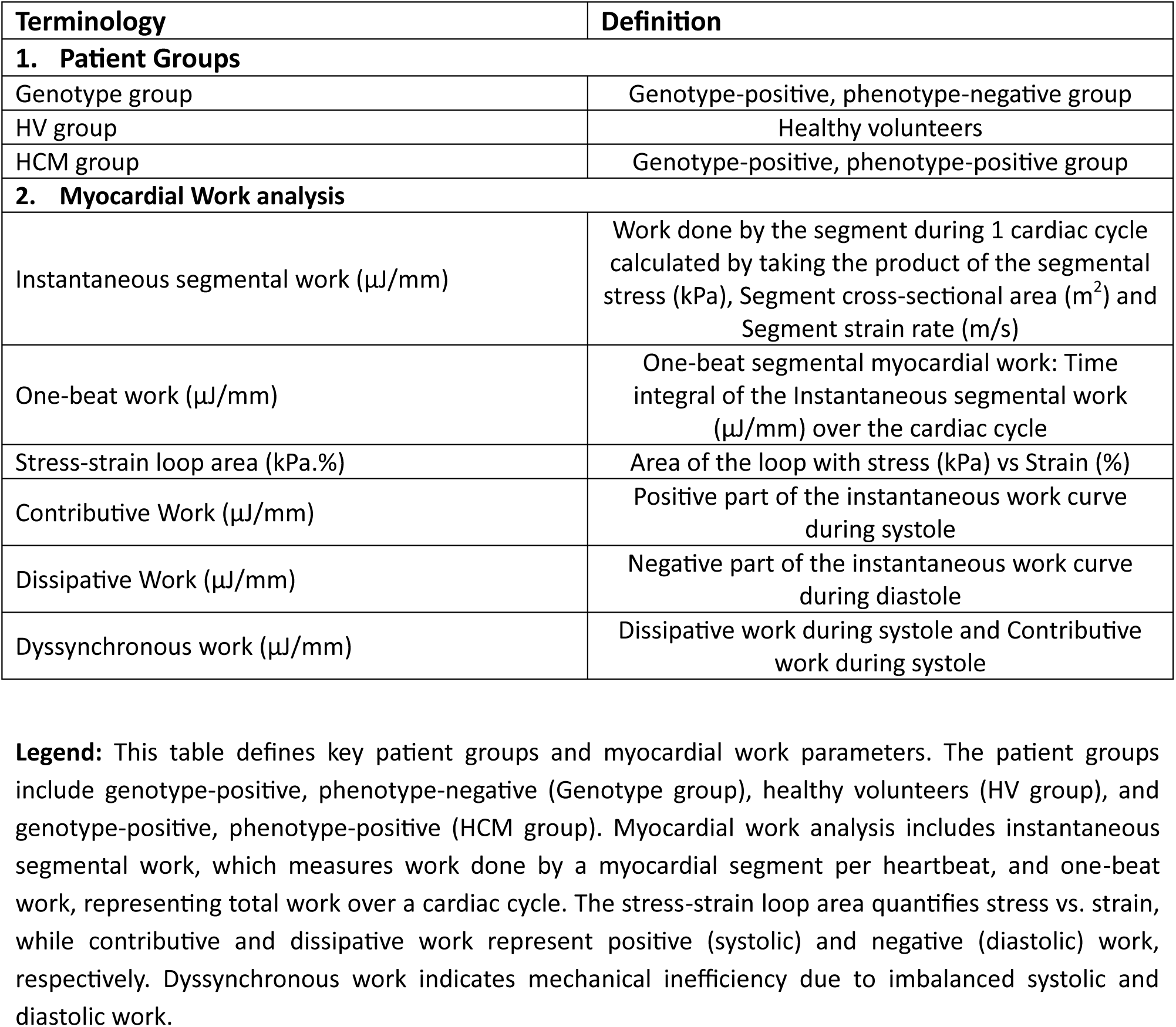
Terminologies and definitions of the patient groups and work analyses.

| Terminology | Definition |
| --- | --- |
| **1. Patient Groups** |  |
| Genotype group | Genotype-positive, phenotype-negative group |
| HV group | Healthy volunteers |
| HCM group | Genotype-positive, phenotype-positive group |
| **2. Myocardial Work analysis** |  |
| Instantaneous segmental work ($\mu\text{J}/\text{mm}$) | Work done by the segment during 1 cardiac cycle calculated by taking the product of the segmental stress (kPa), Segment cross-sectional area ($\text{m}^2$) and Segment strain rate (m/s) |
| One-beat work ($\mu\text{J}/\text{mm}$) | One-beat segmental myocardial work: Time integral of the Instantaneous segmental work ($\mu\text{J}/\text{mm}$) over the cardiac cycle |
| Stress-strain loop area (kPa.%) | Area of the loop with stress (kPa) vs Strain (%) |
| Contributive Work ($\mu\text{J}/\text{mm}$) | Positive part of the instantaneous work curve during systole |
| Dissipative Work ($\mu\text{J}/\text{mm}$) | Negative part of the instantaneous work curve during diastole |
| Dyssynchronous work ($\mu\text{J}/\text{mm}$) | Dissipative work during systole and Contributive work during systole |

**Legend:** This table defines key patient groups and myocardial work parameters. The patient groups include genotype-positive, phenotype-negative (Genotype group), healthy volunteers (HV group), and genotype-positive, phenotype-positive (HCM group). Myocardial work analysis includes instantaneous segmental work, which measures work done by a myocardial segment per heartbeat, and one-beat work, representing total work over a cardiac cycle. The stress-strain loop area quantifies stress vs. strain, while contributive and dissipative work represent positive (systolic) and negative (diastolic) work, respectively. Dyssynchronous work indicates mechanical inefficiency due to imbalanced systolic and diastolic work.

The stress-strain loop area (kPa·%) quantifies the mechanical work performed by the myocardium at the fiber level, integrating myocardial stress (force per unit area) and strain (deformation) over the cardiac cycle. This measurement provides insight into myocardial efficiency and energy expenditure. Both one-beat work and the stress-strain loop area are computed from pressure-strain relationships and are depicted in **Figure 1**.

In addition to the two time-averaged variables, we analyze the instant value of work throughout the cardiac cycle (**Figure 1**). By convention, the axes orientations are selected so that a positive work corresponds to segment contraction. Consequently, the positive part of the instant work curve represents the active contraction of the segment during systole and is termed “contributive work”, as it contributes to the ejection of the heart. Conversely, the negative part of the instant work curve, occurring physiologically in early diastole, represents the work expended by the segment that does not contribute to its shortening, as the resultant forces lead to segment lengthening. This negative work is termed “dissipative work” and can be viewed as the energy expended to resist the stretching of the segment. Instances where dissipative work occurs during systole or contributive work during diastole are considered as “desynchronized work”. The timing of aortic valve closure (AVC) was determined from the cine-echo loop and utilized to distinguish between systole and diastole using the same time scale based on the ECG segmentation described below. Different components of work computed in this study are summarized in Table 1.

### Myocardial Stress-Strain loops

Using ECG correlation, temporal loops were obtained combining myocardial stress and strain results. Area of this loop was extracted using a semi-automatized graphic user interface, custom-created in MATLAB software (California, USA).

### Statistical analysis

The normality of the distribution of the quantitative variables of interest was established with the Shapiro Wilks test. Descriptive statistics were used for quantitative variables. To measure the central tendency we used means, for variation we used ranges and standard deviation. For categorical variables proportions were used. For the inferential statistics, the Student’s T test was used for scalar variables of independent samples. To determine correlation, Pearson coefficient was obtained for normally distributed data and Spearman Coefficient for non-normally distributed data. The Chi^2^ test was used to compare proportions of dichotomous categorical variables. A value of p<0.05 was considered significant. One-way ANOVA was used to compare the quantitative variables for multiple groups. All statistical analyses were performed using MATLAB (version R2020b) and PRISM (2022).

## RESULTS

### Population Characteristics

An age-matched population of 20 HV (mean age = 11.1 ± 4.5 years), 20 HCM (mean age 11.6 ± 5.3 years), and 20 Genotype patients (mean age = 11.1 ± 4.8 years) were included in the study (total = 60 participants). All HCM patients had predominantly septal hypertrophy, 7/20 (35%) HCM patients were under cardiac medications, and all these patients were taking b-blockers. One HCM patient was also taking amiodarone and one HCM patient was also taking calcium blockers. Of the 20 patients in the HCM group, 10 had a MYH7 mutation and 10 had a MYBPC3 mutation. In the Genotype group (n=20) we had patients with the following genotypes: 10 MYBPC3, 4 MYH7, 2 TNNT2, 1 TNNC1, 1 TNNI3, 1 TTN and 1 TPM1.

The demographic data of the population study are summarized in Table 2. There is no significant difference in the weight and body surface area of the 3 groups (p> 0.05, see Table 2).

**Table 2.**
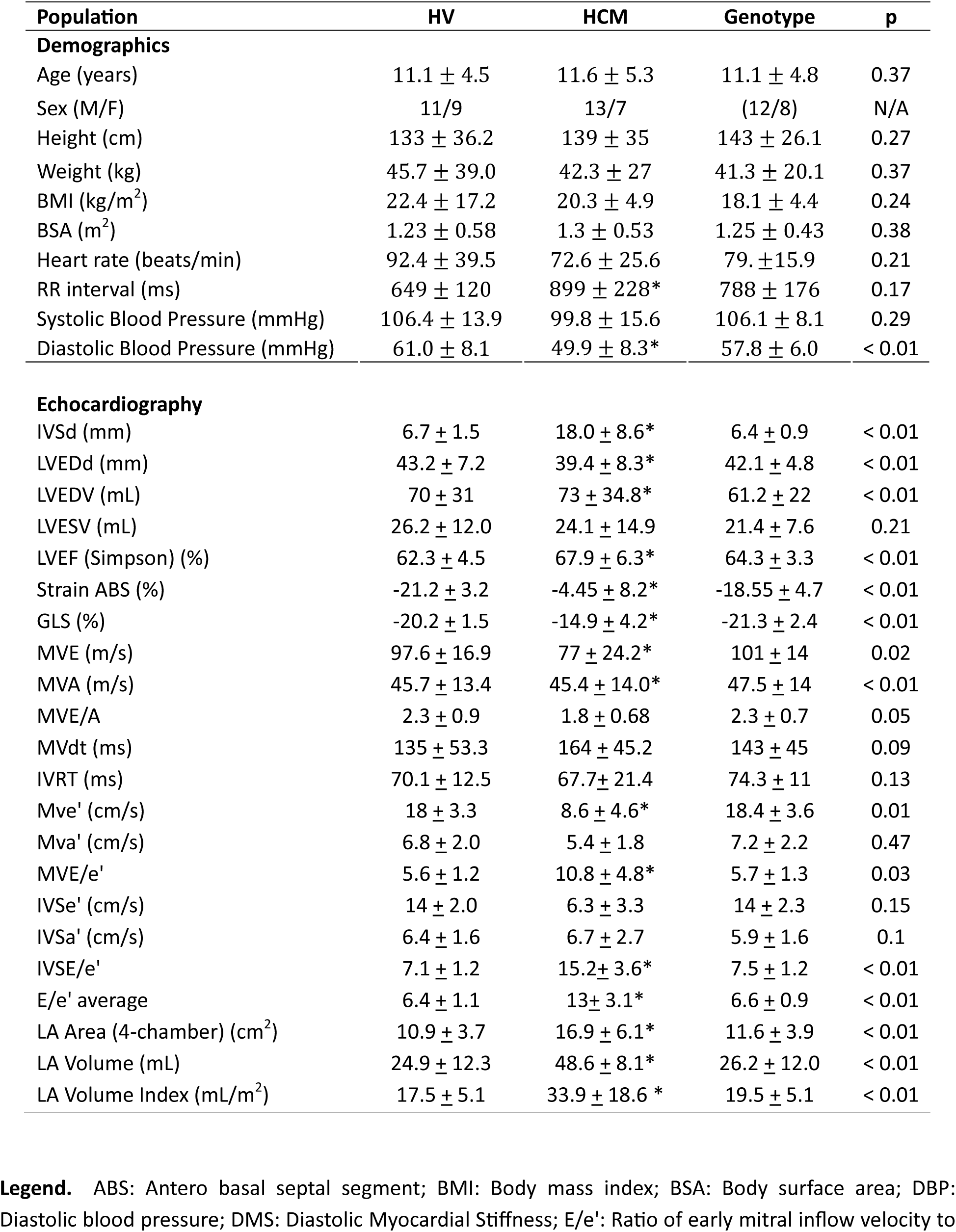
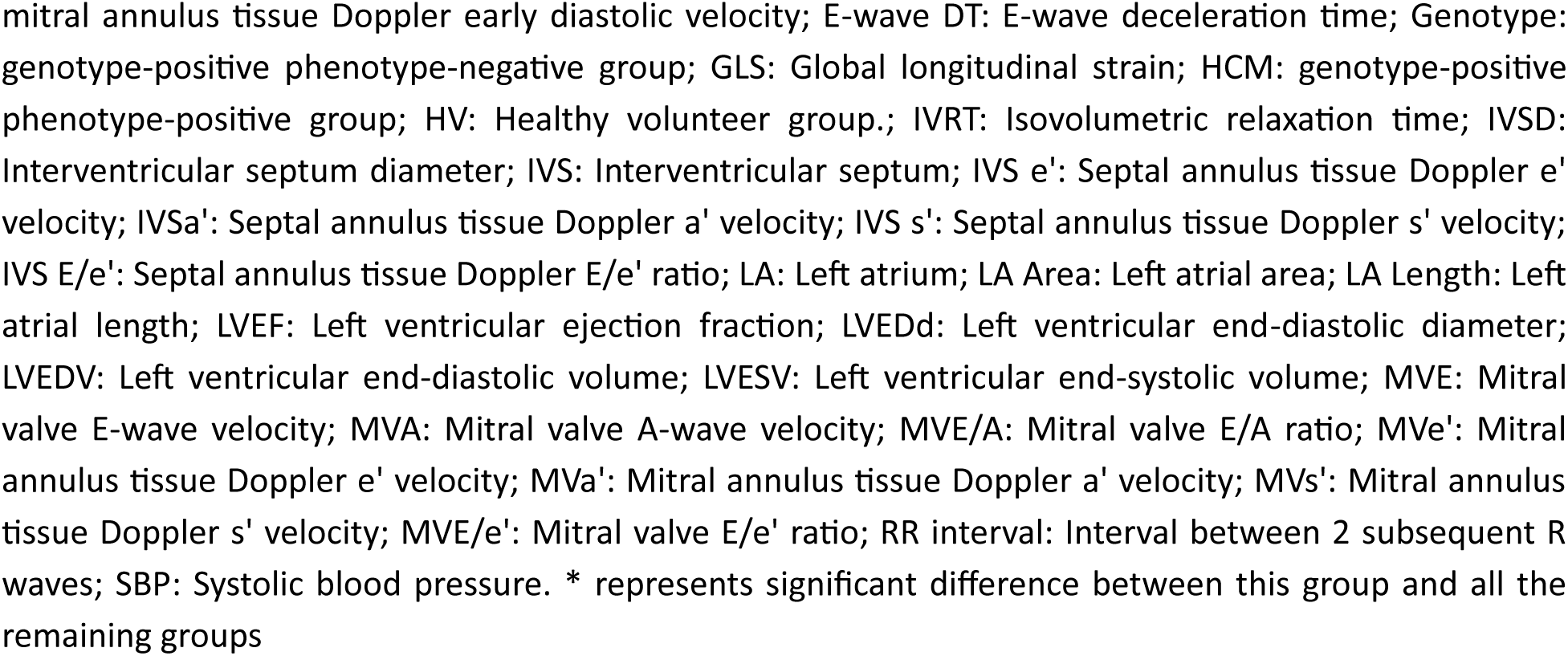
Demographics and Echocardiography data.

| Population | HV | HCM | Genotype | p |
| --- | --- | --- | --- | --- |
| **Demographics** |  |  |  |  |
| Age (years) | 11.1 ± 4.5 | 11.6 ± 5.3 | 11.1 ± 4.8 | 0.37 |
| Sex (M/F) | 11/9 | 13/7 | (12/8) | N/A |
| Height (cm) | 133 ± 36.2 | 139 ± 35 | 143 ± 26.1 | 0.27 |
| Weight (kg) | 45.7 ± 39.0 | 42.3 ± 27 | 41.3 ± 20.1 | 0.37 |
| BMI (kg/m<sup>2</sup>) | 22.4 ± 17.2 | 20.3 ± 4.9 | 18.1 ± 4.4 | 0.24 |
| BSA (m<sup>2</sup>) | 1.23 ± 0.58 | 1.3 ± 0.53 | 1.25 ± 0.43 | 0.38 |
| Heart rate (beats/min) | 92.4 ± 39.5 | 72.6 ± 25.6 | 79. ± 15.9 | 0.21 |
| RR interval (ms) | 649 ± 120 | 899 ± 228* | 788 ± 176 | 0.17 |
| Systolic Blood Pressure (mmHg) | 106.4 ± 13.9 | 99.8 ± 15.6 | 106.1 ± 8.1 | 0.29 |
| Diastolic Blood Pressure (mmHg) | 61.0 ± 8.1 | 49.9 ± 8.3* | 57.8 ± 6.0 | < 0.01 |
| **Echocardiography** |  |  |  |  |
| IVSd (mm) | 6.7 ± 1.5 | 18.0 ± 8.6* | 6.4 ± 0.9 | < 0.01 |
| LVEDd (mm) | 43.2 ± 7.2 | 39.4 ± 8.3* | 42.1 ± 4.8 | < 0.01 |
| LVEDV (mL) | 70 ± 31 | 73 ± 34.8* | 61.2 ± 22 | < 0.01 |
| LVESV (mL) | 26.2 ± 12.0 | 24.1 ± 14.9 | 21.4 ± 7.6 | 0.21 |
| LVEF (Simpson) (%) | 62.3 ± 4.5 | 67.9 ± 6.3* | 64.3 ± 3.3 | < 0.01 |
| Strain ABS (%) | -21.2 ± 3.2 | -4.45 ± 8.2* | -18.55 ± 4.7 | < 0.01 |
| GLS (%) | -20.2 ± 1.5 | -14.9 ± 4.2* | -21.3 ± 2.4 | < 0.01 |
| MVE (m/s) | 97.6 ± 16.9 | 77 ± 24.2* | 101 ± 14 | 0.02 |
| MVA (m/s) | 45.7 ± 13.4 | 45.4 ± 14.0* | 47.5 ± 14 | < 0.01 |
| MVE/A | 2.3 ± 0.9 | 1.8 ± 0.68 | 2.3 ± 0.7 | 0.05 |
| MVdt (ms) | 135 ± 53.3 | 164 ± 45.2 | 143 ± 45 | 0.09 |
| IVRT (ms) | 70.1 ± 12.5 | 67.7 ± 21.4 | 74.3 ± 11 | 0.13 |
| Mve' (cm/s) | 18 ± 3.3 | 8.6 ± 4.6* | 18.4 ± 3.6 | 0.01 |
| Mva' (cm/s) | 6.8 ± 2.0 | 5.4 ± 1.8 | 7.2 ± 2.2 | 0.47 |
| MVE/e' | 5.6 ± 1.2 | 10.8 ± 4.8* | 5.7 ± 1.3 | 0.03 |
| IVSe' (cm/s) | 14 ± 2.0 | 6.3 ± 3.3 | 14 ± 2.3 | 0.15 |
| IVSa' (cm/s) | 6.4 ± 1.6 | 6.7 ± 2.7 | 5.9 ± 1.6 | 0.1 |
| IVSE/e' | 7.1 ± 1.2 | 15.2 ± 3.6* | 7.5 ± 1.2 | < 0.01 |
| E/e' average | 6.4 ± 1.1 | 13 ± 3.1* | 6.6 ± 0.9 | < 0.01 |
| LA Area (4-chamber) (cm<sup>2</sup>) | 10.9 ± 3.7 | 16.9 ± 6.1* | 11.6 ± 3.9 | < 0.01 |
| LA Volume (mL) | 24.9 ± 12.3 | 48.6 ± 8.1* | 26.2 ± 12.0 | < 0.01 |
| LA Volume Index (mL/m<sup>2</sup>) | 17.5 ± 5.1 | 33.9 ± 18.6 * | 19.5 ± 5.1 | < 0.01 |

**Legend.** ABS: Antero basal septal segment; BMI: Body mass index; BSA: Body surface area; DBP: Diastolic blood pressure; DMS: Diastolic Myocardial Stiffness; E/e': Ratio of early mitral inflow velocity to
mitral annulus tissue Doppler early diastolic velocity; E-wave DT: E-wave deceleration time; Genotype: genotype-positive phenotype-negative group; GLS: Global longitudinal strain; HCM: genotype-positive phenotype-positive group; HV: Healthy volunteer group.; IVRT: Isovolumetric relaxation time; IVSD: Interventricular septum diameter; IVS: Interventricular septum; IVS e': Septal annulus tissue Doppler e' velocity; IVSa': Septal annulus tissue Doppler a' velocity; IVS s': Septal annulus tissue Doppler s' velocity; IVS E/e': Septal annulus tissue Doppler E/e' ratio; LA: Left atrium; LA Area: Left atrial area; LA Length: Left atrial length; LVEF: Left ventricular ejection fraction; LVEDd: Left ventricular end-diastolic diameter; LVEDV: Left ventricular end-diastolic volume; LVESV: Left ventricular end-systolic volume; MVE: Mitral valve E-wave velocity; MVA: Mitral valve A-wave velocity; MVE/A: Mitral valve E/A ratio; MVe': Mitral annulus tissue Doppler e' velocity; MVa': Mitral annulus tissue Doppler a' velocity; MVs': Mitral annulus tissue Doppler s' velocity; MVE/e': Mitral valve E/e' ratio; RR interval: Interval between 2 subsequent R waves; SBP: Systolic blood pressure. \* represents significant difference between this group and all the remaining groups

### Conventional Echocardiography

Echocardiography results are summarized in Table 2.

### Myocardial stiffness and strain

The variations throughout the cardiac cycle of myocardial stiffness, myocardial strain, myocardial strain rate and wall thickness of the basal antero-septal segment are presented in Figure 2. The results for myocardial stiffness (Figure 2a) show that there is no significant difference between the peak systolic stiffness between the HVs (47.7 ± 3.8 kPa, SWV = 3.8 ± 0.43 m/s), HCM (60.7 ± 22.8 kPa, SWV = 4.5 ± 0.35 m/s, p = 0.43) or Genotype (39.3 ± 6.9 kPa, SWV = 3.5 ± 0.32m/s) groups, (p = 0.13). The mean diastolic myocardial stiffness (DMS) is significantly higher in the HCM group (23.7 ± 8.7 kPa, SWV = 2.9 ± 0.28 m/s) compared to the HVs (7.2 ± 0.7 kPa, SWV = 1.55 ± 0.36 m/s), p<0.01. However, there is no significant difference between the mean DMS between the HVs and the Genotype group (7.93 ± 1.1 kPa, SWV = 1.62 ± 0.89 m/s), (p value= 0.57).

**Figure 2.**
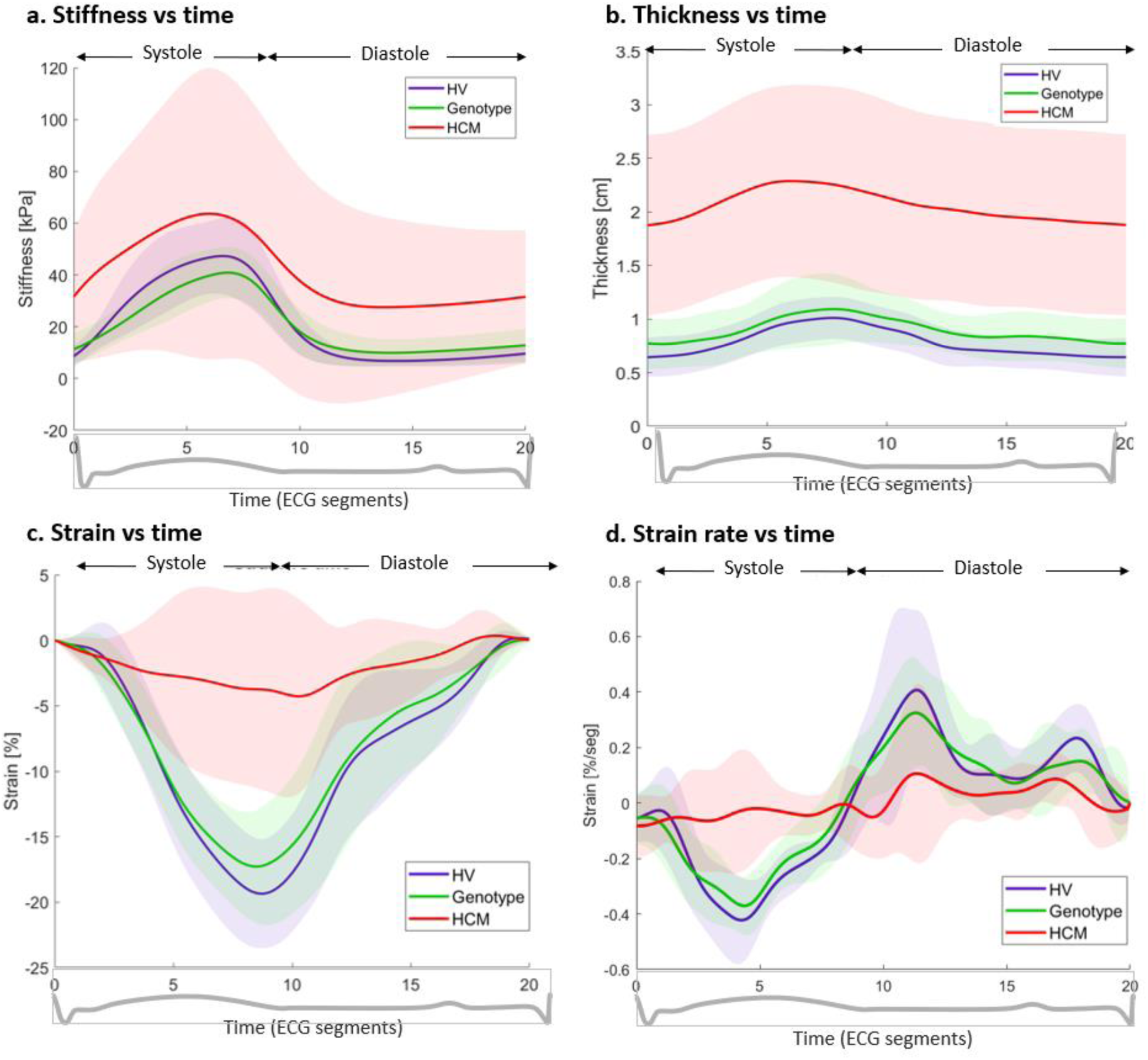
Myocardial stiffness, thickness, strain, and strain rate measurements throughout the cardiac cycle. **Legend**. (a) Myocardial stiffness of the basal antero-septal segment. (b) Wall thickness of the basal antero-septal segment. (c) Longitudinal myocardial strain of the basal antero-septal segment. (d) Strain rate of the basal antero-septal segment. Genotype: genotype-positive phenotype-negative group; HCM: genotype-positive phenotype-positive group; HV: healthy volunteer group.

As shown in Figure 2c, the peak systolic strain also shows no significant difference between the HVs (−19.9 ± 6.1%) and the genotype group (−16.8 ± 3.6%) (p = 0.41). However, the HCM group (−6.64 ± 8.9%) has significantly lower peak systolic strain compared to the HVs (−19.9 ± 4.1%), p<0.01. Similar results are observed for strain rate among the 3 groups shown in Figure 2d (HV = −0.466 ± 0.14 %/segment, HCM = - 0.27 ± 0.15 %/segment, Genotype = −0.402 ± 0.08 %/segment).

### Myocardial stress

As illustrated in Figure 3, the peak systolic myocardial stress is similar between the HVs (−8.72 ± 3.7 kPa) and Genotype group (−6.43 ± 2.2 kPa, p=0.1). The HCM group (−4.25 ± 4.5 kPa) has significantly lower peak systolic myocardial stress compared to HVs (−8.72 ± 3.7 kPa) (p <0.01) as shown in Figure 3b.

**Figure 3.**
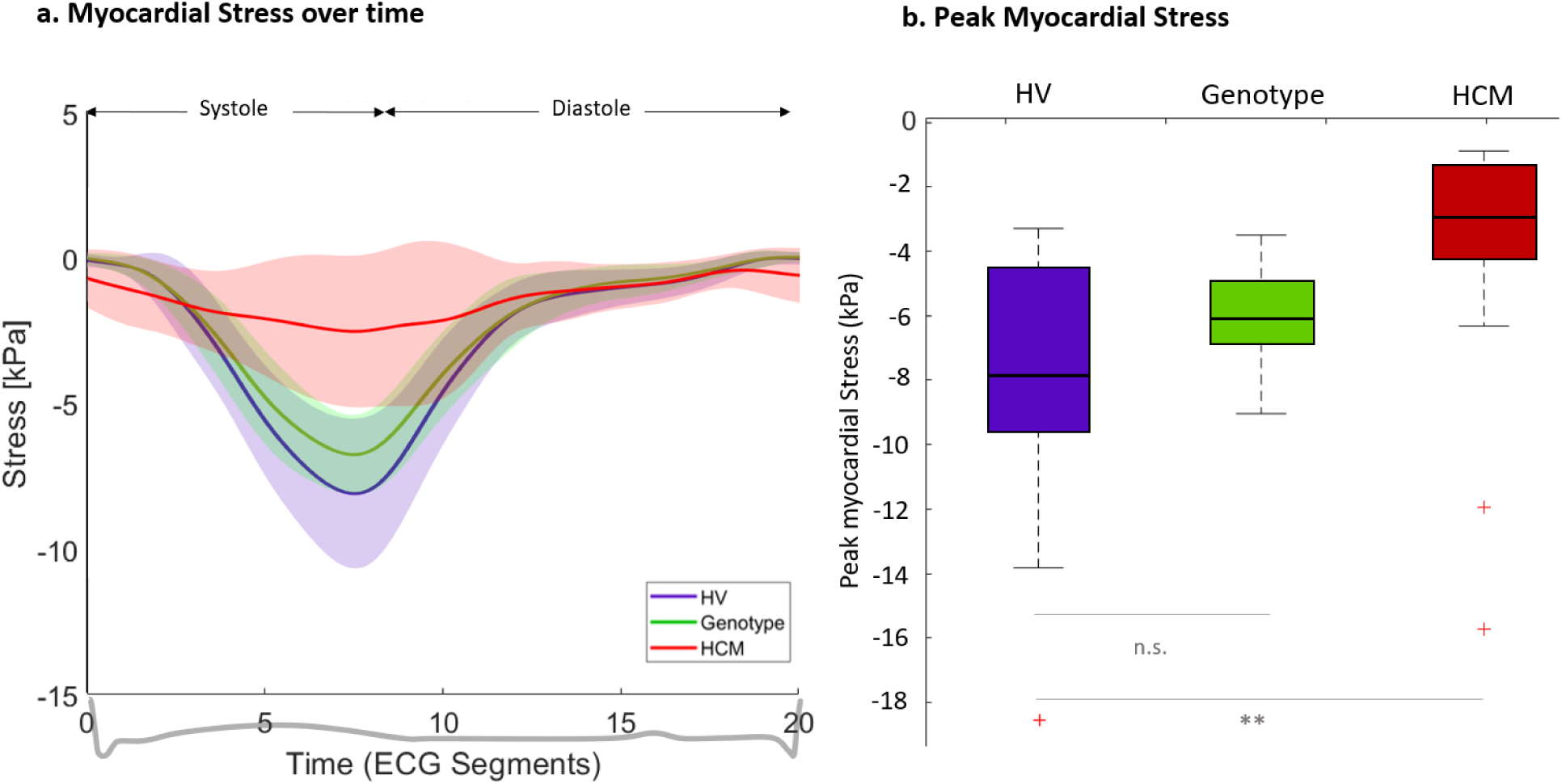
Myocardial stress over the cardiac cycle. **Legend**. (a) Myocardial stress is presented over time. (b) Peak myocardial stress for the 3 groups. Genotype: genotype-positive phenotype-negative group; HCM: genotype-positive phenotype-positive group; HV: healthy volunteer group.

### Myocardial work

Figure 4a. shows the variation in instantaneous segmental work throughout the cardiac cycle for all groups. Figure 4b illustrates the comparison of one-beat work, contributive work, and dissipative work across the three study groups. One-beat work was significantly lower in the HCM and Genotype groups compared to HVs (p<0.01). Similarly, contributive work was reduced in both HCM and Genotype groups relative to HVs (p<0.01). However, dissipative work remained comparable across all three groups (p=0.41). Complete numerical results can be found in Figure 4b or Supplemental Table 1.

**Figure 4.**
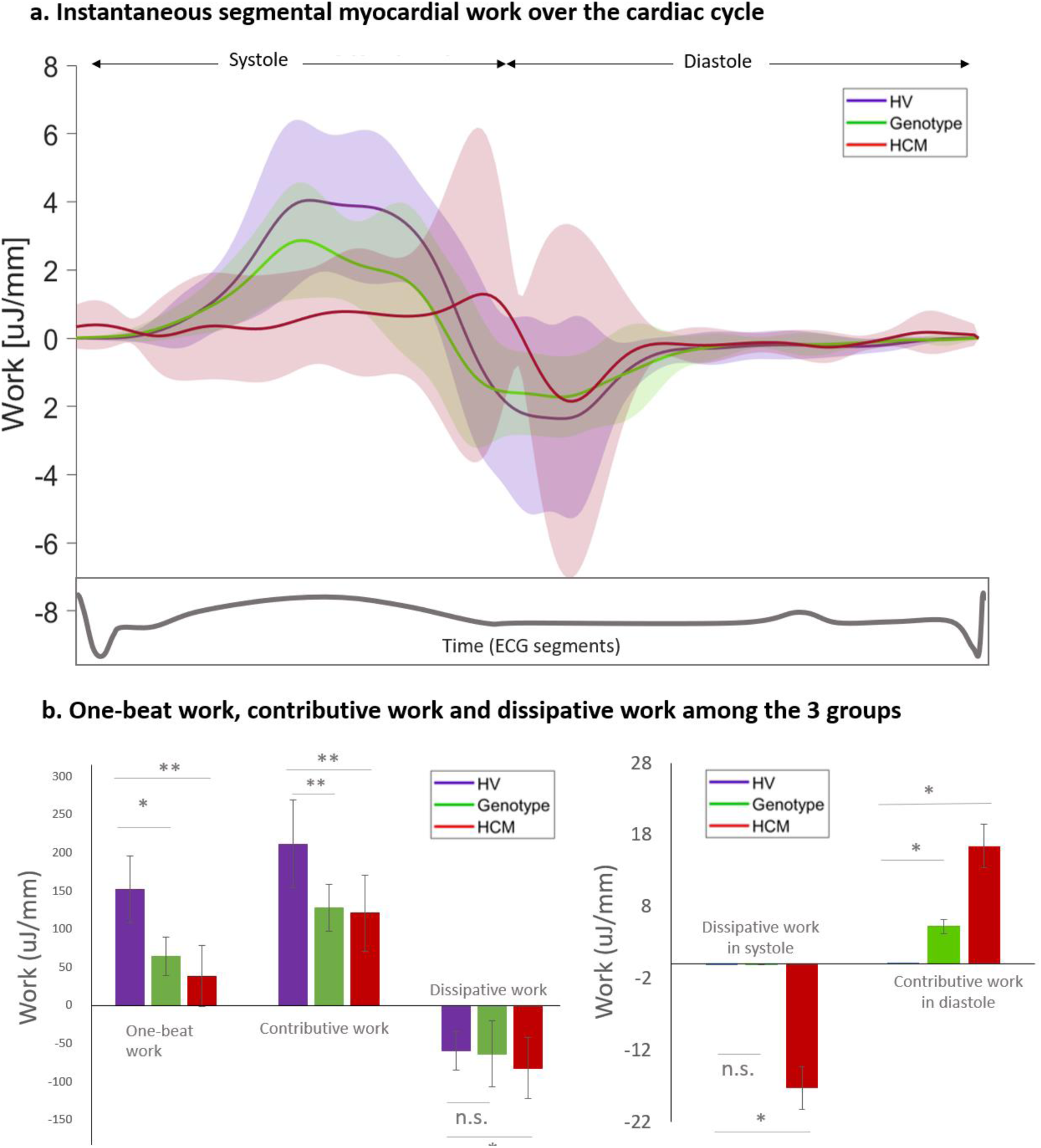
Instantaneous and One-beat work analysis over the full cardiac cycle. **Legend:** Panel a: Variation in instantaneous segmental work during the cardiac cycle is shown for the 3 groups. Panel b: One-beat, contributive and dissipative work is presented for the three groups (left) along with the desynchronized dissipative work during systole and contributive work during diastole (right). Genotype: genotype-positive phenotype-negative group; HCM: genotype-positive phenotype-positive group; HV: healthy volunteer group; One-beat work: One-beat segmental myocardial work.

Dyssynchronous work during systole and diastole was observed for HCM patients but was also observed for Genotype group during diastole (Figure 5b. right panel). For the HCM group, dyssynchronous work was quantified during systole as dissipative work of 17.3 ± 28.9 µJ/mm (45% of one-beat work) and 15.3 ± 18.0 µJ/mm of contributive work during diastole (40% of the one-beat work). For the Genotype group, dyssynchronous work was quantified as 5.6 ± 3.2 µJ/mm of contributive work during diastole (5% of the one-beat work).

**Figure 5.**
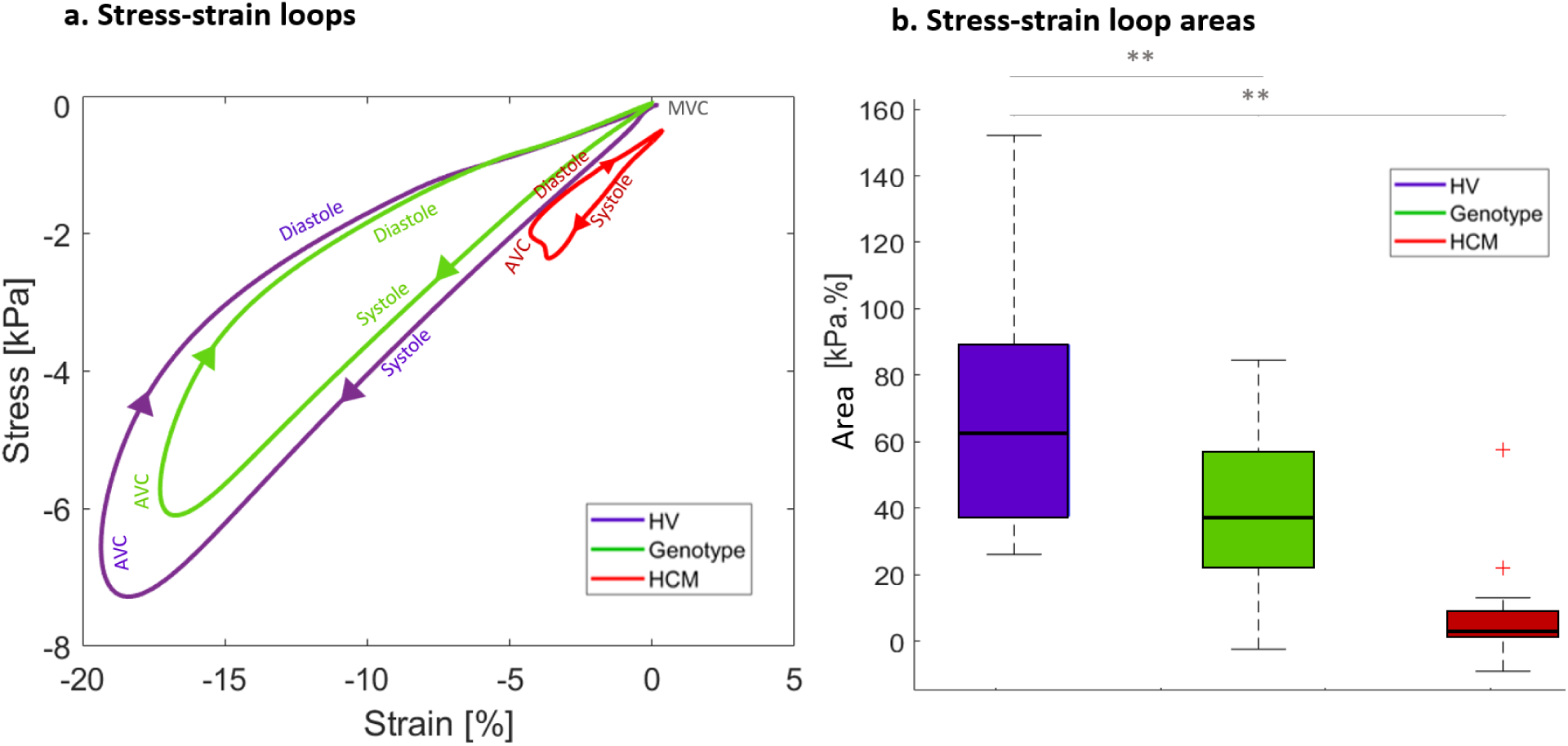
Stress-strain loops. **Legend** (a) Myocardial stress strain loops presented for the 3 groups. (b) Comparison of the stress-strain loop area for the 3 groups. AVC: aortic valve closure; AVO: aortic valve opening; Genotype: genotype-positive phenotype-negative group; HCM: genotype-positive phenotype-positive group; HV: healthy volunteer group; MVC: mitral valve closure; MVO: mitral valve opening; MS: myocardial stiffness N.S: no significant difference.

### Myocardial Stress-Strain loops

The stress vs strain loops for the 3 groups are shown in Figure 5. The stress-strain loop area is significantly lower in HCM group (5.8 ± 6.5 kPa. %) as compared to the HVs (66.8 ± 19.8 kPa. %), p <0.01. The Genotype group has significantly lower (33.2 ± 10.6 kPa. %) stress-strain loop area compared to the HVs (66.8 ± 19.8 kPa. %), p value< 0.01 but significantly higher than the HCM group (5.8 ± 6.5 kPa. %), p value< 0.01.

### Genotype Specific Analysis

Comparison of diastolic myocardial stiffness, one-beat work and stress-stress loop area among the specific genotypes of the HCM group and Genotype group are presented in Table 3. The results show no significant difference in the comparison of these specific genotypes for myocardial stiffness, one-beat work and stress-stress loop area.

**Table 3.** Comparison of genotype specific results.

|  | HCM group |  |  | Genotype group |  |  |
| --- | --- | --- | --- | --- | --- | --- |
| Parameter | MYH7 | MYBPC3 | p | Thick myofilament mutations | Thin myofilament mutations | p |
| **Diastolic Stiffness (kPa)** | 2.42 ± 1.7 | 3.15 ± 1.2 | 0.13 | 1.85 ± 0.4 | 1.98 ± 0.5 | 0.3 |
| **One Beat Work (μJ/mm)** | 23.3 ± 4.2 | 29.1 ± 8.6 | 0.41 | 47.9 ± 9.2 | 51.8 ± 8.6 | 0.47 |
| **Stress-strain loop area (kPa. %)** | 5.1 ± 1.2 | 5.3 ± 3.1 | 0.78 | 39.5 ± 3.6 | 43.1 ± 3.6 | 0.56 |

**Legend.** Genotype: genotype-positive phenotype-negative group; HCM: genotype-positive phenotype-positive group; HV: healthy volunteer group; One-beat work: One-beat segmental myocardial work; Thick myofilament mutations: MYH7 and MYBPC3; and Thin myofilament mutations: TNNT2, TNNC1, TNNI3, TTN, TPM1.

## DISCUSSION

In the present investigation, we demonstrated that individual echocardiographic parameters such as myocardial thickness, strain, strain rate, myocardial stiffness, and stress values, do not differ between the Genotype group and healthy controls, while all these parameters are significantly reduced in the phenotypical HCM group. Our results show that using a combination of myocardial stress and strain, along with dynamic myocardial thickness, derived parameters such as one-beat work and stress-strain loop areas, differ in both Genotype and HCM groups in comparison to HVs. Our results suggests that the combined parameters could be valuable for more in-depth phenotypical characterization. In patients with the phenotype, these parameters may provide additional information to aid in predicting outcomes. Furthermore, this highlights the complexity of myocardial mechanics in Genotype patients, where subclinical myocardial changes may not be adequately detected using traditional parameters such as myocardial thickness or strain alone.

### Myocardial stress and strain: non-invasive assessment of myocardial work

Traditionally, quantification of myocardial work has relied on invasive measures, primarily the pressure-volume loop area, which provides major information about myocardial and cardiac performances. Several studies have highlighted the role of myocardial work (MW) in hypertrophic cardiomyopathy (HCM) and its association with fibrosis, microvascular dysfunction, and disease progression. Jacquemy et al. (2024) demonstrated that impaired myocardial work in children with HCM correlates with left ventricular fibrosis, suggesting MW as a potential marker of disease severity [19]. Similarly, García Brás et al. (2023) linked microvascular dysfunction to impaired MW in both obstructive and non-obstructive HCM, highlighting its role in disease heterogeneity [20].

Further, Galli et al. (2019) showed that constructive MW is reduced in HCM patients and predicts left ventricular fibrosis, reinforcing its diagnostic significance [21]. Additionally, Hiemstra et al. (2020) demonstrated that altered MW in non-obstructive HCM patients has implications for risk stratification and prognosis [22]. These studies underscore the clinical relevance of MW assessment in HCM and support its potential use as a non-invasive tool for patient monitoring and outcome prediction.

In recent years, non-invasive methods using pressure-deformation loop area have emerged which exhibit linear correlations between myocardial work and oxygen consumption, offering clinically viable alternatives that enhance patient comfort and accessibility for the assessment of cardiac work. Russell et al. (2012) presented a novel clinical method for quantifying left ventricular pressure-strain loop area, highlighting its potential as a non-invasive index of myocardial work [23]. Our proposed method includes the use of myocardial strain, which has been widely adopted in clinical practice due to its sensitivity for detecting early subclinical myocardial dysfunction. It is also considered a standard parameter for left ventricular (LV) quantification in guideline recommendations [24]. However, strain values can vary significantly depending on preload and afterload, making it challenging to establish definitive thresholds for pathological changes [25]. In contrast, myocardial work allows us to integrate the geometric variations of the septal segment throughout the cardiac cycle, providing a more accurate assessment of myocardial function by reflecting both deformation and the forces that influence it. This is particularly important in the context of hypertrophic cardiomyopathy, where wall thickness and myocardial strain are critical factors in disease progression. [1]

Contrary to ventricular pressure estimates used in previous works, our method used myocardial stress over the full cardiac cycle which captures the full spectrum of intrinsic constraint imposed on the myocardium [26], [27]. Therefore, integrating stress and thickness measurements adds an additional layer of information about the segmental shortening and lengthening of myocardial segments. This combination enables the evaluation of myocardial work along time —positive work during systole and negative work during diastole—providing a more complete understanding of the dynamic functionality of the myocardium throughout the cardiac cycle. This integrated approach captures the full mechanical performance of myocardial segments at each time point, offering a more precise and comprehensive evaluation of cardiac function in HCM patients [26], [28].

### Clinical implications and phenotypic Variability

From a clinical perspective, our results identify stress-strain loop areas, along with one-beat work, as parameters showing significant differences in the Genotype patients (i.e. with known sarcomeric gene mutation but without LV hypertrophy) from HVs—despite the Genotype group being clinically considered normal due to normal septal thickness, as per the American College of Cardiology/American Heart Association guidelines on pediatric HCM [1]. While our results have not highlighted a specific genotype associated with abnormal myocardial performance (stress, work, or loops), several studies have highlighted the substantial variability in phenotypic expression, even among individuals with the same genetic mutation [8], [29]. The incomplete penetrance of HCM-associated genetic mutations underscores the multifactorial nature of the disease and advancement in cardiac imaging has now become important for the integration of structural and functional information in addition to molecular analysis to provide the basis of new therapeutic interventions [8], [30].

### Myocardial segmental work and desynchrony

Previous studies done by our team has demonstrated myocardial stiffness as a new potential parameter for the assessment of diastolic dysfunction in a pediatric population, both for healthy volunteers and HCM patients [11], [14], [18]. In our recent study we demonstrated the importance of considering ventricular geometry when assessing myocardial stiffness, particularly myocardial thickness for the HCM population. Subsequently, to see how the viscoelastic property of the myocardial tissue translates towards its dynamic function of actively contracting and relaxing during systole and diastole, we combine myocardial stiffness with myocardial strain and segmental thickness to compute myocardial stress and subsequently one-beat work using Hook’s law. The results of our study show a significant reduction in one-beat work for HCM patients compared to HVs, which is consistent with the results of similar studies that use pressure-strain loops for work assessment to estimate myocardial fibrosis in HCM patients [21]. Furthermore, we were able to quantify a significant degree of dyssynchronous work during systole and diastole for the HCM patients. Dyssynchronous contraction, in the absence of intra/interventricular conduction defects on the ECG, is common in patients with HCM, especially if they have significant septal hypertrophy [33], [34]. However, it was interesting to observe a small degree of dyssynchronous work during diastole for the Genotype group which may have the potential to be a new parameter for risk stratification for this “normal” phenotype population [35]. Our current findings suggest a potential role in phenotype risk stratification; however, further clinical validation is needed to confirm these observations and their correlation with clinical outcomes.

### Limitations

An important limitation of this study was the relatively small sample size of 20 subjects per group which made it challenging to compare results for specific genotypes. Further studies with large cohort numbers could help correlate results with specific gene mutations. Additionally, the assessment was performed for only the antero-basal septal segment which limits our ability to generalize results for other segments of the myocardium. Furthermore, we were unable to obtain pressure-volume loop area measurements for comparison with our results due to the highly invasive procedure of cardiac catheterization. Lastly from a methodological perspective, the myocardial strain assessment does not take into consideration the changes in these parameters along the circumferential tensor [15], [36].

## CONCLUSION

In conclusion, we demonstrated a novel non-invasive approach to estimating One-beat segmental myocardial work (one-beat work) by combining myocardial stiffness, strain, and thickness throughout the cardiac cycle. Our findings suggest that one-beat work and stress-strain loops have the potential distinguish Genotype patients from healthy volunteers. While it is not likely to replace myocardial strain, it may add incremental value to existing strain evaluation in HCM patients, offering a more accurate and individualized assessment of myocardial function. Further studies are needed to determine correlations between these new mechanical myocardial performance metrics and clinical outcomes in HCM or Genotype patients.

## Data Availability

All data referenced and analyzed in this manuscript are available from the corresponding author upon reasonable request. All datasets generated during the current study are archived in secure institutional storage and will be shared in de-identified form for academic research purposes.

## Acknowledgements

No

## ABBREVIATIONS

DMS: diastolic myocardial stiffness
HCM: hypertrophic cardiomyopathy
IVRT: isovolumetric relaxation time
MVC: mitral valve closure
MS: myocardial stiffness
MW: myocardial work
PMS: peak myocardial strain
STE: speckle tracking echocardiography
SWE: shear wave elastography
SWV: shear wave velocity
UUI: ultrafast ultrasound imaging

---

**Supplemental Table 1.** Myocardial work measurements.

| Work Type | HV Group | Genotype Group | HCM Group |
| --- | --- | --- | --- |
| One-Beat Work ($\mu\text{J}/\text{mm}$) | $152.1 \pm 44.2$ | $64.0 \pm 25.1$ | $38.2 \pm 107.1$ |
| Contributive Work ($\mu\text{J}/\text{mm}$) | $211.7 \pm 58.1$ | $127.6 \pm 30.4$ | $120.9 \pm 134$ |
| Dissipative Work ($\mu\text{J}/\text{mm}$) | $-59.6 \pm 24.8$ | $-63.6 \pm 43.5$ | $-82.6 \pm 83.8$ |
| Dyssynchronous Work - Systole ($\mu\text{J}/\text{mm}$) | $< 0.01$ | $< 0.01$ | $17.3 \pm 28.9$ (45% of one-beat work) |
| Dyssynchronous Work – Diastole ($\mu\text{J}/\text{mm}$) | $< 0.01$ | $5.6 \pm 3.2$ (5% of one-beat work) | $15.3 \pm 18.0$ (40% of one-beat work) |

**Legend.** This table presents the comparative values of one-beat work, contributive work, dissipative work, and dyssynchronous work for three study groups: Healthy Volunteers, Genotype, and HCM patients. Genotype: genotype-positive phenotype-negative group; HCM: genotype-positive phenotype-positive group; HV: healthy volunteer group and One-beat work: One-beat segmental myocardial work.

